# Multi-steroid profiling by uPLC-MS/MS with post-column infusion of ammonium fluoride

**DOI:** 10.1101/2022.03.08.22271681

**Authors:** Lina Schiffer, Fozia Shaheen, Lorna C. Gilligan, Karl-Heinz Storbeck, James Hawley, Brian G. Keevil, Wiebke Arlt, Angela E. Taylor

## Abstract

**Background:** Multi-steroid profiling is a powerful analytical tool that simultaneously quantifies steroids from different biosynthetic pathways. Here we present an ultra-high performance liquid chromatography-tandem mass spectrometry (uPLC-MS/MS) assay for the profiling of 25 steroids using post-column infusion of ammonium fluoride.

**Methods:** Following liquid-liquid extraction, steroids were chromatographically separated over 5 minutes using a Phenomenex Luna Omega C_18_ column and a water (0.1 % formic acid) methanol gradient. Quantification was performed on a Waters Acquity uPLC and Xevo^®^ TQ-XS mass spectrometer. Ammonium fluoride (6 mmol/L, post-column infusion) and formic acid (0.1 % (vol/vol), mobile phase additive) were compared as additives to aid ionisation.

**Results:** Post-column infusion (PCI) of ammonium fluoride (NH_4_F) enhanced ionisation in a steroid structure-dependent fashion compared to formic acid (122-140% for 3βOH-Δ5 steroids and 477-1274% for 3-keto-Δ4 steroids). Therefore, we fully analytically validated PCI with NH_4_F. Lower limits of quantification ranged from 0.28 to 3.42 nmol/L; 23 of 25 analytes were quantifiable with acceptable accuracy (bias range −14% to 11.9%). Average recovery ranged from 91.6% to 113.6% and average matrix effects from −29.9% to 19.9%. Imprecision ranged from 2.3% to 23.9% for all analytes and was <15% for 18/25 analytes. The serum multi-steroid profile of 10 healthy men and 10 healthy women was measured.

**Conclusions:** uPLC-MS/MS with post-column infusion of ammonium fluoride enables comprehensive multi-steroid profiling through enhanced ionisation particularly benefiting the detection of 3-keto-Δ4 steroids.

**Highlights:**

- This multi-steroid profiling assay quantifies 25 steroids in 5.5 minutes
- Post-column infusion of NH_4_F enhances the ionisation of 3-keto-Δ4 steroids
- The assay simultaneously quantifies steroids from several biosynthetic pathways
- We present analytical data validated for serum steroid profiling

## 1. Introduction

Steroid hormones are biosynthesised in the adrenal cortex and gonads via cascade-like, interlinked enzymatic pathways and undergo extensive metabolism with both activation and inactivation in peripheral tissues (Figure 1) leading to a complex circulating steroid metabolome [1]. Steroid flux through the different pathways can be severely dysregulated in various conditions, requiring comprehensive steroid assessment to develop a mechanistic understanding of the condition and for diagnosis and treatment monitoring. Disorders of steroidogenesis with distinct steroid metabolome profiles include inborn enzymatic deficiencies such as congenital adrenal hyperplasia, autonomous adrenal steroid production such as Cushing syndrome or primary aldosteronism [2], adrenocortical carcinoma [3, 4], polycystic ovary syndrome and idiopathic intracranial hypertension [5-7]. Steroid-dependent cancers, such as prostate cancer, can locally activate steroids and are treated with various pharmacological or surgical approaches to deplete the relevant steroids and their precursors in circulation [8].

**Figure 1:**
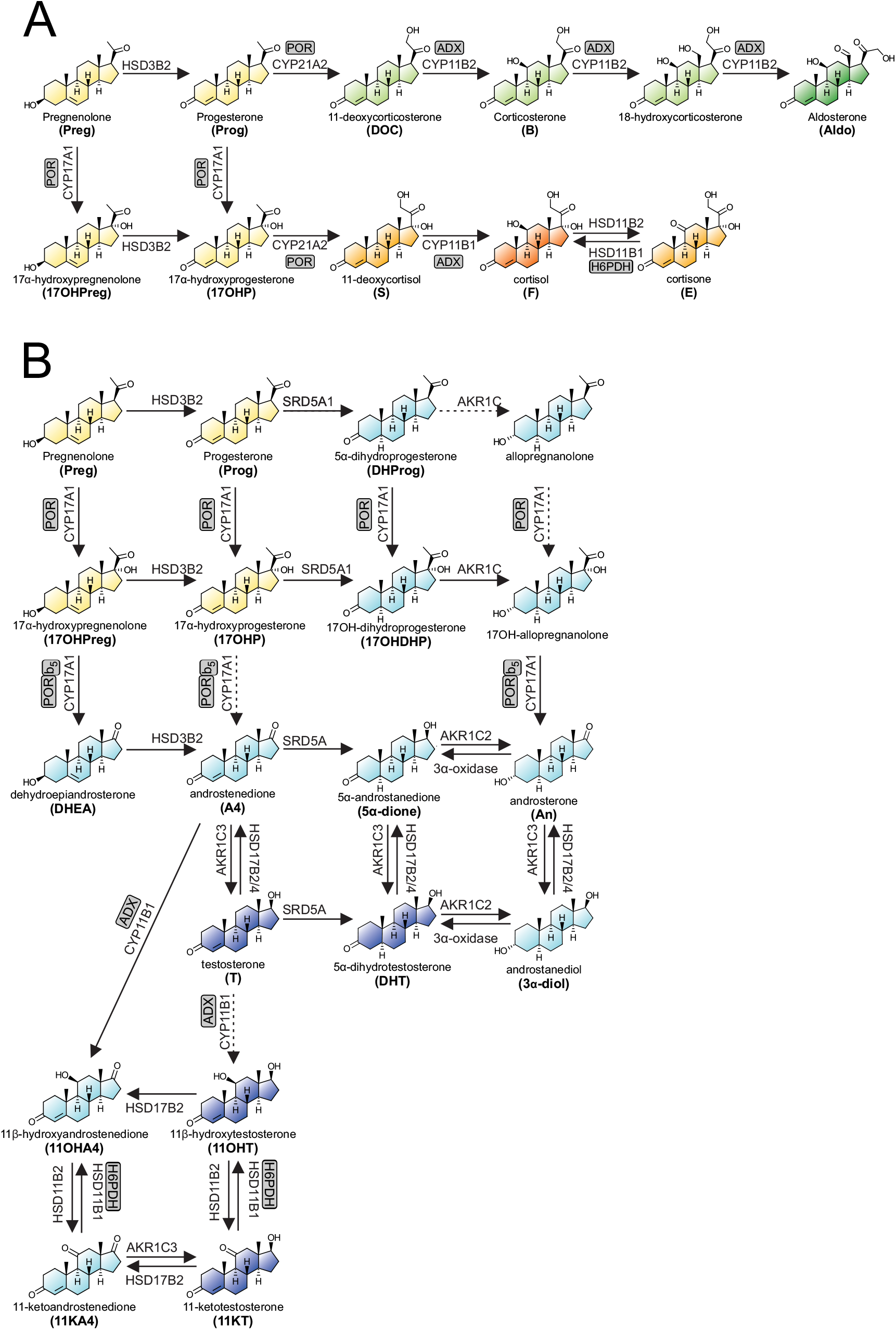
Pathways of adrenal steroidogenesis (A) and androgen biosynthesis (B). General steroid precursors are shown in yellow, mineralocorticoids in green, glucocorticoids in orange, androgens in blue. Dark shades of each colour indicate active steroids, light shades inactive precursors. Abbreviations for steroids included in the uPLC-MS/MS assay are shown in bold.

Traditionally, individual steroids have been used for the assessment of a suspected underlying condition (e.g. testosterone (T) for female androgen excess [9]; 17α-hydroxyprogesterone (17OHP) for congenital adrenal hyperplasia [10]). Mechanistic in-vitro studies often rely on the measurement of the end products of a biosynthesis pathway only. However, the determination of a multi-steroid profile has several advantages over the use of selected individual markers. First, steroid precursor/product ratios can be calculated to identify distinct effects on individual enzymes [11] and to assess the degree of steroid precursor activation in conditions of steroid excess. For example, in serum the ratio of T to its less active precursor androstenedione (A4) and the ratio of T to the more potent androgen 5α-dihydrotestosterone (DHT) have been shown to be excellent markers for androgen excess and the associated adverse metabolic phenotype in polycystic ovary syndrome [5, 12, 13]. Secondly, multi-steroid profiling enables different pathways of steroidogenesis to be assessed simultaneously, which is essential due to the interlinked nature of steroidogenesis and the contribution of individual enzymes to several pathways (Figure 1). Thirdly, multi-steroid profiling can be used to investigate off-target effects of inhibitors of steroidogenesis. Finally, multi-steroid profiling can be combined with machine learning approaches to generate powerful, unbiased and automated diagnostic steroid metabolomics tools [3, 4].

In contrast to immunoassays, mass spectrometry allows for multiplexing of analytes and liquid chromatography for the high-throughput that is required to effectively use multi-steroid profiling in clinical and research laboratories. Steroid analysis by mass spectrometry is often limited by poor sensitivity due to low analyte concentrations in biological samples and low ionisation efficiency of the analytes. Mobile phase additives can improve the chromatography (peak separation and shape), enhance the signal and, thereby, sensitivity. Formic (methanoic) acid and acetic acid are common additives for corticosteroids and androgen analysis in the positive ionisation mode [5, 14, 15] as they can promote the formation of protonated molecular ions (M+H)^+^. Ammonium fluoride (NH_4_F) can aid the ionisation of steroids in electrospray ionisation (ESI) in negative mode [16] and hence improve the sensitivity of oestrogen measurements [17, 18]. Additionally, NH_4_F has been reported to augment the ionisation of steroids using ESI in positive ion mode in a structure-dependent manner, when coupled to supercritical fluid chromatography [19]. Here, we present an ultra-high performance liquid chromatography-tandem mass spectrometry (uPLC-MS/MS) assay to measure 25 steroids (Table 1) from the mineralocorticoid, glucocorticoid, androgen, alternative and 11-oxygenated steroidogenic pathways. The separation was achieved in 5 minutes using post-column infusion of NH_4_F in positive ionisation mode to enhance ionisation.

**Table 1:**
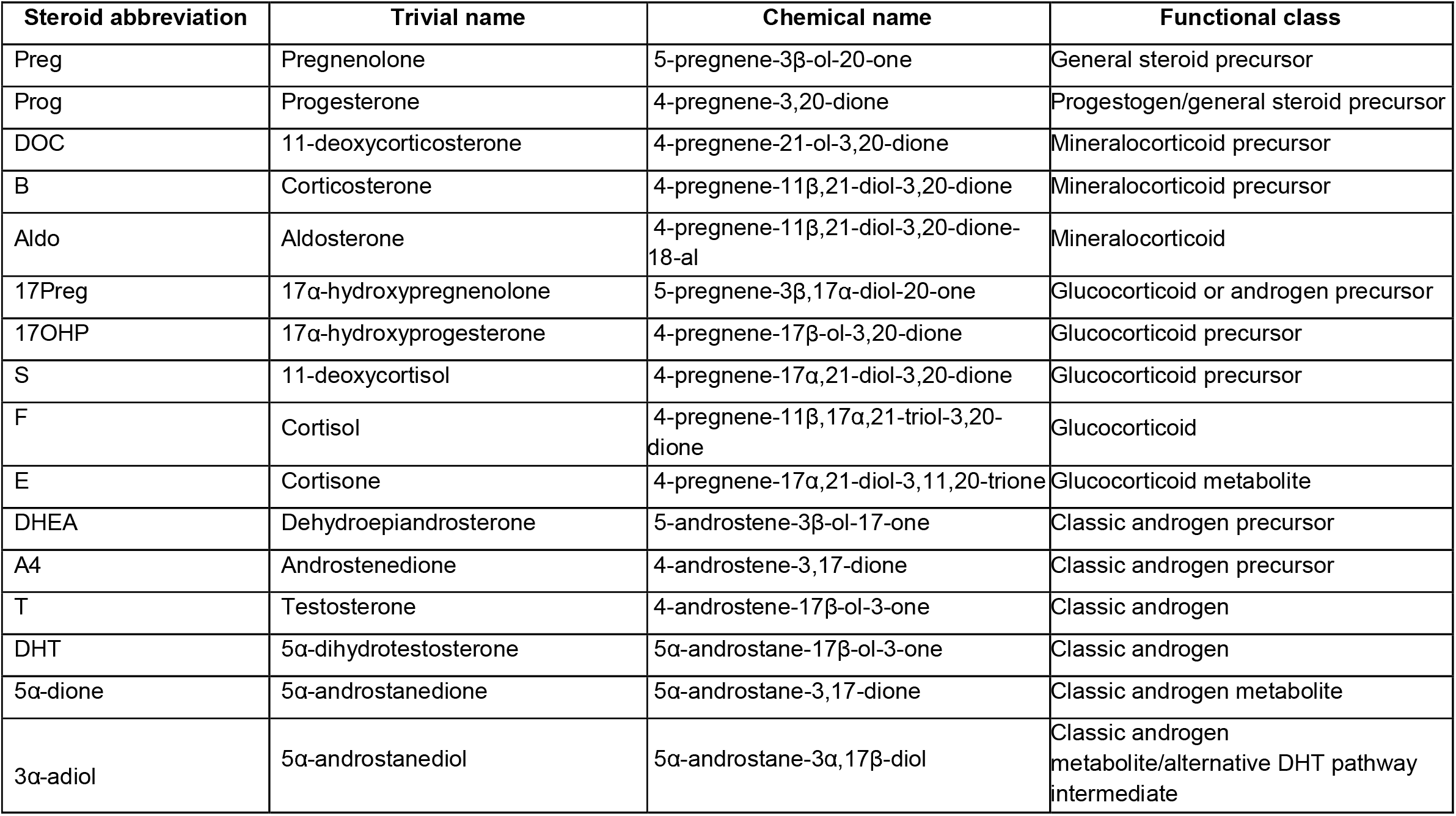
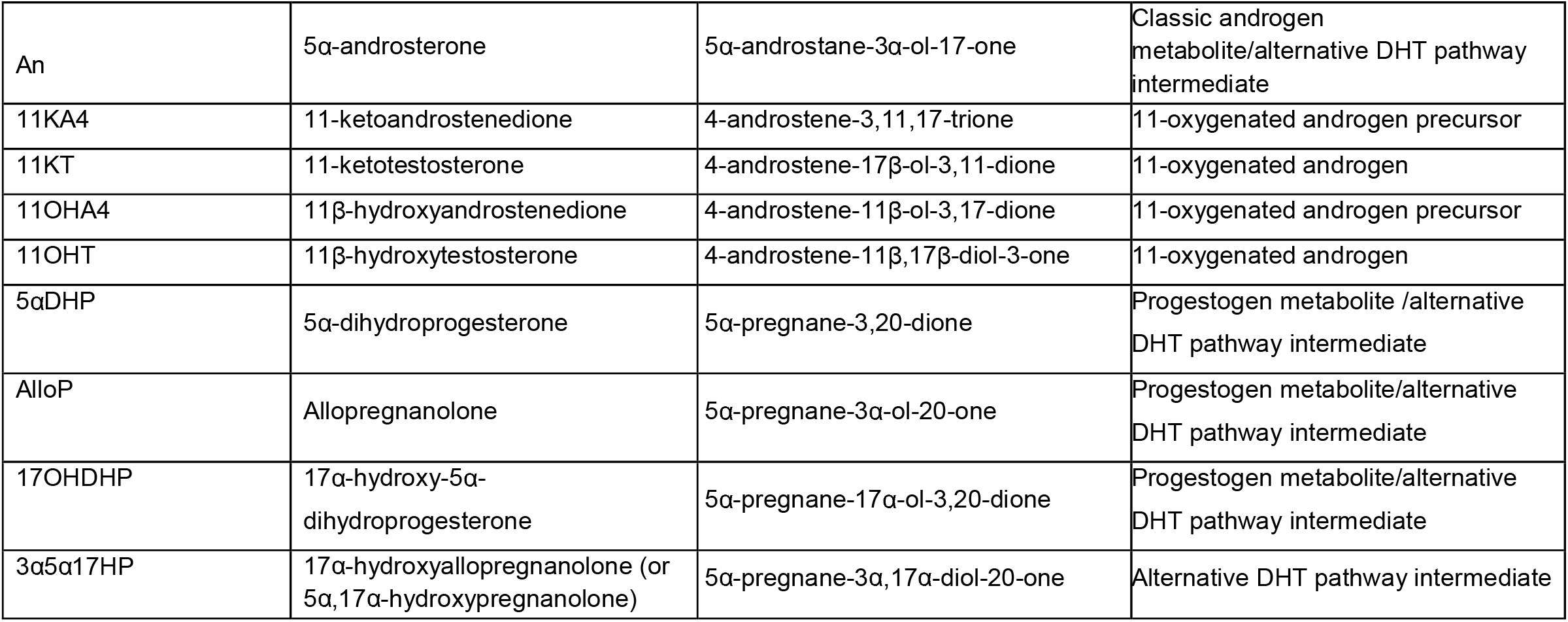
Nomenclature of the 25 steroids analysed in this uPLC-MS/MS assay

| Steroid abbreviation | Trivial name | Chemical name | Functional class |
| --- | --- | --- | --- |
| Preg | Pregnenolone | 5-pregnene-3β-ol-20-one | General steroid precursor |
| Prog | Progesterone | 4-pregnene-3,20-dione | Progestogen/general steroid precursor |
| DOC | 11-deoxycorticosterone | 4-pregnene-21-ol-3,20-dione | Mineralocorticoid precursor |
| B | Corticosterone | 4-pregnene-11β,21-diol-3,20-dione | Mineralocorticoid precursor |
| Aldo | Aldosterone | 4-pregnene-11β,21-diol-3,20-dione-18-al | Mineralocorticoid |
| 17Preg | 17α-hydroxypregnenolone | 5-pregnene-3β,17α-diol-20-one | Glucocorticoid or androgen precursor |
| 17OHP | 17α-hydroxyprogesterone | 4-pregnene-17β-ol-3,20-dione | Glucocorticoid precursor |
| S | 11-deoxycortisol | 4-pregnene-17α,21-diol-3,20-dione | Glucocorticoid precursor |
| F | Cortisol | 4-pregnene-11β,17α,21-triol-3,20-dione | Glucocorticoid |
| E | Cortisone | 4-pregnene-17α,21-diol-3,11,20-trione | Glucocorticoid metabolite |
| DHEA | Dehydroepiandrosterone | 5-androstene-3β-ol-17-one | Classic androgen precursor |
| A4 | Androstenedione | 4-androstene-3,17-dione | Classic androgen precursor |
| T | Testosterone | 4-androstene-17β-ol-3-one | Classic androgen |
| DHT | 5α-dihydrotestosterone | 5α-androstane-17β-ol-3-one | Classic androgen |
| 5α-dione | 5α-androstanedione | 5α-androstane-3,17-dione | Classic androgen metabolite |
| 3α-adiol | 5α-androstenediol | 5α-androstane-3α,17β-diol | Classic androgen metabolite/alternative DHT pathway intermediate |
| An | 5α-androsterone | 5α-androstane-3α-ol-17-one | Classic androgen metabolite/alternative DHT pathway intermediate |
| 11KA4 | 11-ketoandrostenedione | 4-androstene-3,11,17-trione | 11-oxygenated androgen precursor |
| 11KT | 11-ketotestosterone | 4-androstene-17β-ol-3,11-dione | 11-oxygenated androgen |
| 11OHA4 | 11β-hydroxyandrostenedione | 4-androstene-11β-ol-3,17-dione | 11-oxygenated androgen precursor |
| 11OHT | 11β-hydroxytestosterone | 4-androstene-11β,17β-diol-3-one | 11-oxygenated androgen |
| 5αDHP | 5α-dihydroprogesterone | 5α-pregnane-3,20-dione | Progestogen metabolite /alternative DHT pathway intermediate |
| AlloP | Allopregnanolone | 5α-pregnane-3α-ol-20-one | Progestogen metabolite/alternative DHT pathway intermediate |
| 17OHDHP | 17α-hydroxy-5α-dihydroprogesterone | 5α-pregnane-17α-ol-3,20-dione | Progestogen metabolite/alternative DHT pathway intermediate |
| 3α5α17HP | 17α-hydroxyallopregnanolone (or 5α,17α-hydroxypregnanolone) | 5α-pregnane-3α,17α-diol-20-one | Alternative DHT pathway intermediate |

## 2. Material and methods

### 2.1 Preparation of external standards and quality controls

Pure standards of all analytes were purchased as powders from Sigma-Aldrich (Gillingham, UK) and Steraloids (Newport, USA); for details see Supplemental Table 1. The purity of all standards was confirmed by gas chromatography-mass spectrometry. Individual stock solutions at 1 mg/mL were prepared in uPLC grade methanol (Biosolve, Dieuze, France) and stored at −80 °C. Using the stock solutions, combined calibrators for all analytes were prepared by spiking phosphate buffered saline (PBS) pH 7.4 supplemented with 0.1% (wt/vol) bovine serum albumin (BSA) (Sigma-Aldrich) yielding at least 10 different final concentrations from 0 up to 250 ng/mL (Supplemental Table 2). A combined internal standard stock solution containing 0.5 μg/mL of each internal standard was prepared in deuterated methanol (Sigma-Aldrich).

Quality control (QC) samples were prepared by spiking PBS 0.1% BSA from independent 1 mg/mL stock solutions. Four different concentrations covering the expected concentration range human serum (Supplemental Table 2). Additionally, pooled human serum (Sigma-Aldrich) was aliquoted, stored at −80 °C and used as biological QC in each analytical run. All calibrators, internal standards and QCs were stored at −20 °C.

### 2.2 Collection of serum samples

The collection of blood samples was approved by the authors’ Institutional Review Board (Science, Technology, Engineering and Mathematics Ethical Review Committee of the University of Birmingham, ERN_17-0494). Informed, written consent was obtained from all individuals included in this study. Following venous puncture, blood was collected into gold top SST Vacutainers^®^ (Becton, Dickinson, Wokingham, UK). Samples were spun and the serum removed and stored at −80 °C until analysis. The method was applied to 20 serum samples from healthy volunteers 10 female and 10 males (20-40 years).

### 2.3 Sample preparation

Steroids were extracted by liquid-liquid extraction. A set of calibrators, QCs and biological QCs were prepared with each batch of samples. Sample, calibrator, or QC (200 μL) were transferred into a hexamethyldisilazane-treated glass tube and 10 µL of the internal standard mixture containing all stable isotope labelled internal standards listed in Table S1. 50 μL of acetonitrile (Biosolve, Dieuze, FR) was added to precipitate the proteins and the samples were vortexed. 1-mL of tert-butyl methyl ether (Acros Organics, Fisher Scientific UK Ltd, Loughborough, UK) was added to each sample and the samples were vortexed at 1000 rpm for 10 minutes on a multi-vortex. The samples were incubated at room temperature for at least 30 minutes to aid phase separation. The organic phase was subsequently transferred into the wells of a 2-mL square well 96-well plate (Porvair Sciences Ltd, Wrexham, UK) containing 700 µL glass inserts (Randox, Crumlin, UK) and dried under a nitrogen stream at 45 °C. The dried extract was reconstituted in 100 µL of 50% (vol/vol) uPLC grade methanol (Biosolve) in uPLC grade water (Biosolve) prior to analysis.

### 2.4 Ultra-high performance liquid chromatography

Chromatography was performed on an Acquity ultra performance liquid chromatography system (uPLC; Waters Ltd, Wilmslow, UK) using a Phenomenex Luna Omega column, 1.6 µm, polar C18, 100 Å, 2.1 × 50 mm; (Phenomenex, Macclesfield, UK) at 60 °C. 10 µL of the reconstituted sample was injected. Mobile phase A consisted of uPLC grade water (Biosolve) and mobile phase B of uPLC grade methanol (Biosolve). An optimised method with a linear gradient from 45% to 75% of mobile phase B was applied over five minutes at a flow rate of 0.6 mL/min to separate the analytes followed by a 98% wash and equilibration at starting conditions prior to the injection of the next sample. The auto sampler was maintained at 10 °C. To investigate ionisation 0.1% (vol/vol) formic acid (added to mobile phase A or both A and B) was compared to PCI of NH_4_F.

### 2.5 Tandem mass spectrometry

The uPLC eluate was injected into a XEVO^®^ TQ-XS mass spectrometer (Waters Ltd) using ESI in positive ion mode. Post-column infusion 6 mmol/L NH_4_F in 50% (vol/vol) was combined using the fluidics system on the mass spectrometer under full software control. The capillary voltage was maintained at 1.5 kV, the source temperature was 150 °C, desolvation temperature and gas flow were 600 °C and 1200 L/h, cone gas was 150 L/h. MassLynx 4.2 software (Waters Ltd) was used for systems control. Qualifier and quantifier mass transitions, cone voltages and collision energies are summarised in Table 2. TargetLynx software was used for data processing and quantification. Peak area ratios of analyte to internal standard were plotted against the nominal concentrations of the calibrators and 1/x weighting and linear least square regression were used to produce the standard curve.

**Table 2:** Retention times, quantifier and qualifier mass transitions, collision energies and cone voltages of target analytes and internal standards.

| Analyte | Retention time (min) | Mass transition (m/z)<br>Quantifier<br>Qualifier | Cone voltage (V) | Collision energy (eV) |
| --- | --- | --- | --- | --- |
| Preg | 3.65 | 299.2 > 281.1<br>317.1 > 281.1 | 36<br>16 | 12<br>14 |
| Prog | 3.37 | 315.1 > 97.1<br>315.1 > 109.1 | 20<br>20 | 20<br>24 |
| DOC | 2.21 | 331.1 > 97.1<br>331.1 > 109.1 | 26<br>26 | 20<br>24 |
| B | 1.52 | 347.2 > 121.1<br>347.2 > 97.1 | 20<br>16 | 24<br>20 |
| Aldo | 0.86 | 361.2 > 343.2<br>361.2 > 315.1 | 40<br>40 | 16<br>18 |
| 17Preg | 2.42 | 315.2 > 297.2<br>297.2 > 279.2 | 8<br>4 | 12<br>14 |
| 17OHP | 2.45 | 331.1 > 97.1<br>331.1 > 109.1 | 16<br>16 | 22<br>26 |
| S | 1.64 | 347.1 > 109.1<br>347.1 > 97.1 | 42<br>42 | 26<br>22 |
| F | 1.06 | 363.3 > 121.1<br>363.1 > 91.1 | 42<br>42 | 22<br>50 |
| E | 0.96 | 361.1 > 163.1<br>361.1 > 121.01 | 46<br>46 | 22<br>38 |
| DHEA | 2.35 | 271.2 > 253.2<br>289.2 > 253.2 | 30<br>12 | 10<br>8 |
| A4 | 2.03 | 287.2 > 109.1<br>287.2 > 97.1 | 26<br>26 | 22<br>22 |
| T | 2.26 | 289.2 > 97.1<br>289.2 > 109.1 | 40<br>40 | 20<br>24 |
| DHT | 2.80 | 291.2 > 255.1<br>291.2 > 159.1 | 42<br>42 | 14<br>22 |
| 5α-dione | 2.63 | 289.2 > 253.1<br>289.2 > 271.1 | 40<br>40 | 16<br>10 |
| 3α-adiol | 3.21 | 275.2 > 257<br>275.2 > 81 | 28<br>28 | 10<br>28 |
| An | 3.34 | 291.2 > 273.2<br>291.2 > 255.2 | 24<br>24 | 8<br>12 |
| 11KA4 | 1.06 | 301.1 > 121.1<br>301.1 > 257.1 | 44<br>44 | 22<br>22 |
| 11KT | 1.21 | 303.1 > 121<br>303.1 > 259.1 | 20<br>20 | 22<br>22 |
| 11OHA4 | 1.34 | 303.1 > 285.1<br>303.1 > 267.1 | 30<br>30 | 14<br>16 |
| 11OHT | 1.48 | 305.2 > 269.2<br>305.2 > 121.1 | 16<br>16 | 14<br>20 |
| 5αDHP | 4.01 | 317.1 > 281.1<br>317.1 > 85.1 | 38<br>24 | 12<br>14 |
| alloP | 4.33 | 319.1 > 301.2<br>319.1 > 283.2 | 22<br>22 | 8<br>14 |

| Internal standard | Retention time (min) | Mass transition (m/z)<br>Quantifier<br>*Qualifier* | Cone voltage (V) | Collision energy (eV) |
| --- | --- | --- | --- | --- |
| 17OHDHP | 2.95 | 333.2 > 297.2<br>315.1 > 111 | 26<br>32 | 14<br>16 |
| 3α5α17HP | 3.85 | 317.2 > 299.2<br>317.2 > 111 | 28<br>28 | 12<br>18 |
| Preg-d2-<sup>13</sup>C2 | 3.64 | 321.1 > 303.1 | 28 | 8 |
| Prog-d9 | 3.34 | 324.2 > 100.1 | 28 | 24 |
| DOC-d8 | 2.18 | 339.1 > 100.1 | 26 | 24 |
| B-d8 | 1.49 | 355.1 > 337.2 | 46 | 14 |
| Aldo-d8 | 0.85 | 369.2 > 351.1 | 34 | 16 |
| 17Preg-d3 | 2.4 | 336.2 > 300.2 | 12 | 10 |
| 17OHP-d8 | 2.42 | 339.1 > 100.1 | 26 | 24 |
| S-d2 | 1.62 | 349.1 > 97.1 | 42 | 22 |
| F-d4 | 1.06 | 367.2 > 121.1 | 28 | 26 |
| E-d7 | 0.95 | 369.1 > 169.1 | 40 | 24 |
| DHEA-d6 | 2.33 | 277.2 > 219.1 | 22 | 14 |
| A4-d7 | 2.01 | 294.2 > 109.1 | 26 | 22 |
| T-d3 | 2.25 | 292.2 > 97.1 | 46 | 22 |
| DHT-d3 | 2.79 | 294.1 > 258.1 | 22 | 16 |
| 3α-adiol-d3 | 3.20 | 278.2 > 150.2 | 20 | 20 |
| An-d4 | 3.33 | 295.1 > 277.2 | 24 | 12 |
| 11K4-d10 | 1.04 | 311 > 265 | 46 | 16 |
| 11KT-d3 | 1.20 | 306.2 > 262.1 | 20 | 22 |
| 11OHA4-d7 | 1.30 | 310.1 > 292.1 | 30 | 14 |
| 5αDHP-d4 | 3.96 | 323.1 > 305.1 | 22 | 8 |
| alloP-d4 | 4.31 | 323.1 > 305.1 | 22 | 8 |

### 2.6 Validation

Validation was performed following protocols from published guidelines [20-22]. For each steroid, validation has been performed at different concentrations representing low, medium and high concentrations of that steroid in serum. While this method has utility for many applications, we opted to validate for a serum application. We selected serum because it is a complex mixture of steroids, proteins, salts and many other constituents. These constituents can vary between individuals and can each affect steroid extraction and ionisation efficiencies. In addition, serum contains steroids at a range of concentrations, for example low level poor ionising steroids such as DHT (<0.3 ng/mL (1 nM)) and high concentration good ionisers like cortisol (∼18 to 300 ng/mL (50-800 nM)). Optimisation using such a complex matrix means adaptation of this method to other ‘cleaner’ bio-fluids or cell culture supernatants should be simple.

#### 2.6.1 Recovery and matrix effects

Matrix effects and recovery were determined as previously described [23, 24]. Six different human serum samples (four male and two female) were spiked at 5 ng/mL before extraction (pre-extract) and after extraction and reconstitution (post extract). Additionally, the reconstitution solvent (50% (vol/vol) uPLC grade methanol in uPLC grade water) was spiked with the same amounts of analytes (no extract). Percentage matrix effects and recovery were calculated from the concentrations quantified in the samples:

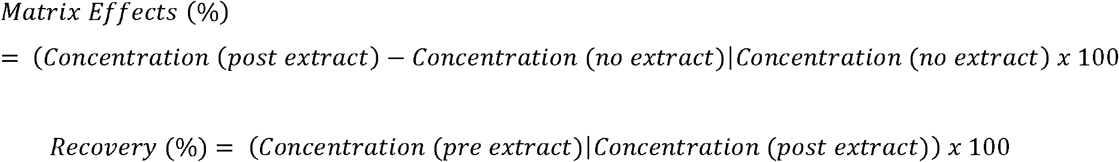

For matrix effects mean values between −15 and 15% were considered ideal and −20% and 20% were acceptable. For recovery mean values between 80% and 120% were defined acceptable.

#### 2.6.2 Linearity

Three calibration series were prepared by different scientists. For each analyte the ratio of analyte peak area to internal standard peak area was plotted against the nominal concentrations of the standard as described above. Calibration curves were accepted as linear if the correlation coefficient of the linear regression (R^2^) was ≥0.99.

#### 2.6.3 Lower limit of quantification

The lower limit of quantification (LLOQ) was defined as the lowest concentration for which 10 replicate samples of spiked surrogate matrix could be run with a CV <20% and a bias not greater than ±20%.

#### 2.6.4 Imprecision

Pooled surrogate matrix samples spiked with all analytes at four different concentrations (0.3, 1, 3 and 30 ng/mL) were run 10 times in the same batch to assess intra-assay imprecision. Pooled serum samples and pooled serum samples spiked with 5 ng/mL for all analytes were also run 10 times in the same batch to assess intra-assay imprecision. These samples were extracted and run-in different batches to assess inter-assay imprecision (n =20). A CV ≤15% was considered optimal.

#### 2.6.5 Accuracy

10 samples of surrogate matrix were spiked individually at four different concentrations (0.3, 1, 3 and 30 ng/mL). A bias of the observed concentration and the nominal concentration between −15% and +15% was considered optimal.

## 3. Results

### 3.1 Chromatographic separation

Analytes were separated using a linear gradient over five minutes followed by 0.5 minutes at starting conditions for equilibration resulting in a total run time injection-to-injection of 5.5 minutes. All analytes eluted as distinct, identifiable peaks (Supplemental Figure 1). Two pairs of analytes co-eluted: (cortisol (*m/z* 363) and 11KA4 (*m/z* 301); 17OHP (*m/z* 330) and 17Preg (*m/z* 332)). No interference was observed due to the differences in *m/z* (Table 2).

### 3.2 Comparison of additives to improve ionisation

The effect of NH_4_F on signal intensity was assessed in comparison to our previously published method using 0.1% (vol/vol) formic acid as mobile phase additive in both the methanol and water phases [5, 7]. We chose to add NH_4_F by post-column infusion as preliminary experiments with NH_4_F as an additive led to increases in system pressure indicative of damage to the column material (data not shown). The intra-assay imprecision using this modification was acceptable with a mean CV of the peak area of 7.0% for all analytes (CV range 3.9 to 13.7%; n=60 injections of the same sample (data not shown)). Peak areas of all analytes in serum (n=73) were compared between the assay with formic acid in both mobile phases and the assay using post-column infusion of NH_4_F. NH_4_F increased peak area in a structure-dependent fashion (Figure 2, Supplemental Figure 2). In comparison to formic acid, NH_4_F induced significant increases in the peak area of steroids with 3-keto-Δ4 structure (Prog, DOC, B, Aldo, 17OHP, S, E, F, A4, 11OHA4, 11KA4, T, 11KT, 11OHT), with mean increases varying from 477% (17OHP) to 1274% (Aldo). NH_4_F had a lower impact on the peak area of the majority of A-ring reduced steroids with changes varying between 100% and 381% (DHT, 5α-dione, 3α-diol, An, 5α-DHP, 3α5α17HP), with the exceptions of alloP (841%) and 17OHDHP (1280%). NH_4_F post-column infusion had only a very minor impact on the peak areas of 3β-OH-Δ5 steroids (Preg, 17Preg, DHEA). Our final optimised method employed 6 mmol/L NH_4_F introduced via post-column infusion at a flow rate of 5 µL/min, with 0.1% (vol/vol) formic acid in the water mobile phase to limit the risk of microbial contamination.

**Figure 2:**
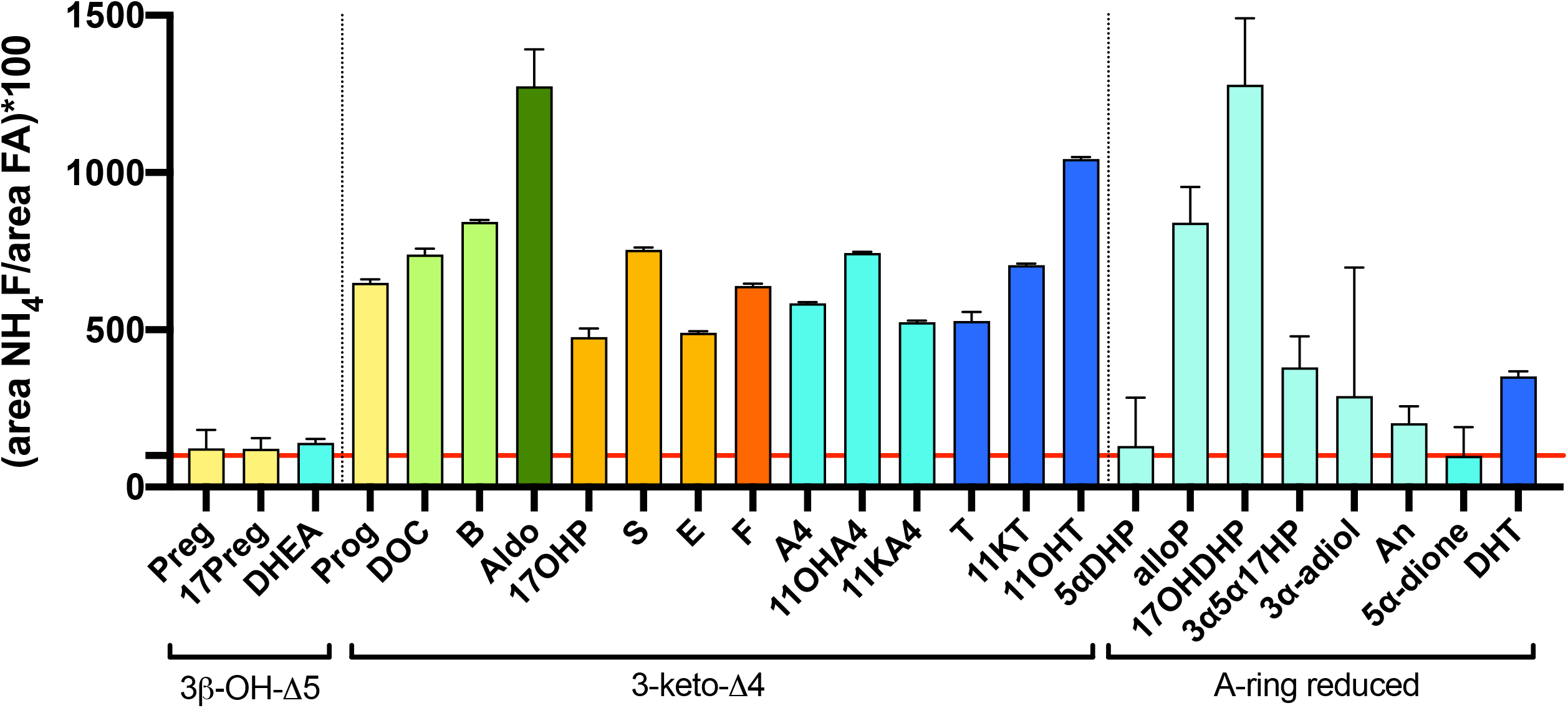
Ammonium fluoride (NH_4_F) post-column infusion enhances ionisation of steroids in a structure-dependent fashion as compared to mobile phase formic acid. The percentage increase in peak area of the quantifier transition when comparing post-column infusion of NH_4_F (6 mmol/L, 5 μL/min) to the use of 0.1% (vol/vol) formic acid in both the methanol and water mobile phase. Bars represent the mean percentage peak area and whiskers the relative standard deviation for serum samples from different individuals. Samples from all individuals with detectable peaks were included (Prog, DOC, B, 17Preg, 17OHP, S, F, E, DHEA, A4, T, DHT, An, 11KA4, 11KT, 11OHA4, 11OHT n=74; Preg n=73; Aldo n=71; 3α-adiol n=56; 5α-DHP n=48; 17OHDHP n=39; 3α5α17HP n=18; alloP n=15; 5α-dione n=14). The red line indicates a relative peak area of 100% (identical ionisation with both methods). The colour of the bars indicates the functional class of each analyte; yellow, general steroid precursor; light green, mineralocorticoid precursor; dark green, mineralocorticoid; orange, glucocorticoid precursor; red, glucocorticoid; turquoise, androgen precursor; dark blue, androgen; light blue, alternative DHT biosynthesis pathway intermediate.

### 3.3 Validation of the analytical performance

The analytical performance of the optimised assay with post-column infusion of NH_4_F and 0.1% (vol/vol) formic acid in mobile phase A was validated. Matrix effects and recovery were assessed for six different serum samples (Table 3). Mean matrix effects ranged from −19.5% to 19.9% for all analytes and were hence in the desired range from −20% to 20%, except for 5αDHP (−29.9%), 17OHDHP (−22.6%) and 3α5α17HP (−62.7%). Mean recovery ranged from 90.0% to 113.6% for all analytes except for 3α5α17HP (122.1%), which was just outside the acceptance range of 80-120%. Calibration curves were linear with an R^2^ ≥0.99 for all analytes (Supplemental Table 2). Lower limits of quantification ranged between 0.1 ng/mL (∼0.3 nmol/L) and 0.5 ng/mL (∼1.5 nmol/L) for all analytes except for 17Preg and 3α-adiol, which had a limit of quantification of 1 ng/mL (corresponding to 3.01 and 3.42 nmol/L, respectively) (Table 3). The accuracy (bias) and imprecision of the assay were assessed with spiked PBS 0.1% BSA samples at four different concentrations. The bias between the observed and nominal concentrations was calculated as a measure of accuracy and the coefficient of variation (CV) as a measure of imprecision (Table 4). Bias was within acceptable limits for the majority of analytes at all concentrations above the LLOQ ranging from −14.0 to 11.9%, with the exception of 5αDHP at 0.3 ng/mL (−21%) and 17OHPreg at 1 ng/mL (−17.0%). Imprecision was within acceptable limits for the majority of analytes at concentrations above the LLOQ ranging from 2.3 to 14.7% with the exception of An (15.2% at 0.3 ng/mL), 3α-adiol (17.8% at 1 ng/mL), 17OHPreg (16.5% at 3 ng/mL) and alloP (at all concentrations imprecision ranged from 15.1 to 20.5%).

**Table 3:**
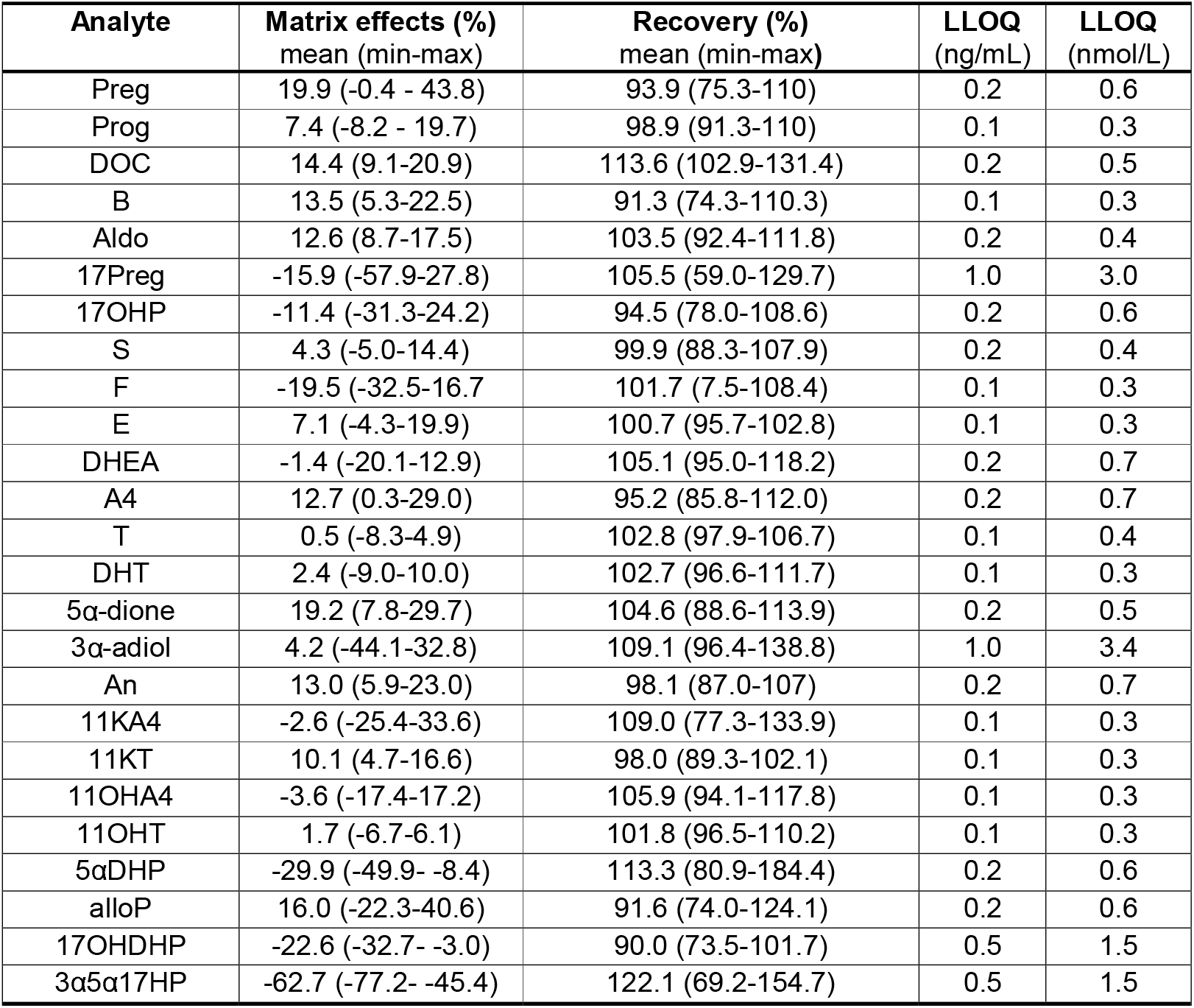
Validation summary of matrix effects, recovery and lower limits of quantification for all steroids. Matrix effects and recovery were assessed at 5 ng/mL (approximately 17 nmol/L) in serum samples from six different donors. The lower limit of quantification (LLOQ) was defined as the lowest concentration that can be assessed with appropriate accuracy (bias within ± 20%) and imprecision (CV < 20%).

**Table 4:** Accuracy and intra-assay imprecision determined at four concentrations spiked into surrogate matrix (n=10). n/a, not applicable as concentration <LLOQ.

| Concentration (ng/mL) | Bias (%) |  |  |  | Imprecision (%) |  |  |  |
| --- | --- | --- | --- | --- | --- | --- | --- | --- |
|  | 0.3 | 1 | 3 | 30 | 0.3 | 1 | 3 | 30 |
| Preg | -9.6 | -0.4 | -1.4 | -3.9 | 8.0 | 5.4 | 13.1 | 4.6 |
| Prog | -6.2 | 1.6 | 1.3 | -5.0 | 7.0 | 5.3 | 9.1 | 3.3 |
| DOC | -0.7 | -2.0 | 3.3 | -0.2 | 7.4 | 7.6 | 11.6 | 6.0 |
| B | -5.4 | -6.5 | -1.8 | 9.2 | 9.4 | 5.8 | 12.1 | 4.8 |
| Aldo | -7.7 | -2.1 | 0.1 | 0.8 | 7.0 | 5.1 | 9.6 | 5.0 |
| 17Preg | n/a | -17 | 3.8 | -3.6 | n/a | 10.4 | 16.5 | 11.1 |
| 17OHP | 4.9 | -0.2 | -5.6 | 0.2 | 5.7 | 10.0 | 9.4 | 6.2 |
| S | 8.4 | 3.9 | 6.8 | 1.8 | 5.5 | 4.8 | 8.3 | 3.8 |
| F | 2.6 | 7.7 | 10.3 | 2.7 | 5.5 | 4.9 | 4.1 | 2.3 |
| E | 0.7 | 4.7 | 7.2 | 5.0 | 6.7 | 4.5 | 8.2 | 4.3 |
| DHEA | 10.6 | 4.8 | 0.6 | 2.3 | 13.3 | 10.1 | 8.0 | 3.8 |
| A4 | 6.1 | -5.1 | -0.4 | -4.6 | 11.3 | 8.2 | 6.9 | 3.7 |
| T | -1.8 | 5.3 | 7.5 | 0.6 | 7.6 | 5.6 | 8.2 | 3.8 |
| DHT | 1.3 | 6.4 | 10.2 | 11.9 | 6.7 | 5.1 | 8.4 | 3.2 |
| 5α-dione | -2.2 | 2.0 | 3.4 | -8.0 | 7.8 | 9.5 | 11.2 | 8.0 |
| 3α-adiol | n/a | 4.4 | -6.1 | 2.5 | n/a | 17.8 | 11.5 | 6.5 |
| An | 2.5 | 1.8 | -0.5 | -2.9 | 15.2 | 12.2 | 9.1 | 3.5 |
| 11KA4 | -9.8 | -5.7 | -0.5 | -0.9 | 8.3 | 6.1 | 11.2 | 5.3 |
| 11KT | -8.5 | 0.5 | 2.3 | -1.6 | 10.5 | 4.8 | 11.6 | 2.7 |
| 11OHA4 | -4.1 | 1.6 | 2.8 | -1.9 | 6.8 | 5.7 | 7.5 | 3.6 |
| 11OHT | 0.2 | 5.6 | 10.9 | 4.0 | 11.6 | 6.8 | 8.0 | 5.0 |
| 5αDHP | -21.0 | -8.2 | -3.4 | -4.6 | 14.3 | 12.5 | 11.6 | 14.4 |
| alloP | -14.0 | -10.7 | -14.0 | -11.5 | 20.5 | 16.1 | 15.7 | 15.1 |
| 17OHDHP | n/a | 4.3 | 9.1 | 6.6 | n/a | 14.7 | 6.5 | 11.6 |
| 3α5α17HP | n/a | 3.5 | 7.5 | 2.0 | n/a | 9.4 | 11.0 | 4.8 |

Additionally, imprecision was assessed using a pooled serum sample (Table 5). The levels in this pooled serum sample were below the limit of quantification for 11 analytes (Preg, Prog, Aldo, DOC, 5α-dione, 3α-adiol, An, 5αDHP, alloP, 17OHDHP, 3α5α17HP). For the remaining analytes the intra-assay CV ranged from 2.4 to 16.3% and the inter-assay CV from 3.5 to 16.1%. For a pooled serum sample spiked with 5 ng/mL of all analytes the intra-assay CV was between 2.3% and 12.5% for all analytes. The inter-assay CV was <15% (3.2 to 14.7%) for all steroids except 17OHDHP (17.2%), 3α-adiol (23.0%), 5α-DHP (22%) and 3α5α17HP (23.9%).

**Table 5:** Intra- and inter-assay precision of a pooled serum sample and a pooled serum sample spiked with 5 ng/mL of all analytes. (n=10). n/a, not applicable as concentration below the lower limit of quantification (<LLOQ).

| Analyte | Pooled serum |  |  | Pooled serum spiked at 5 ng/mL |  |
| --- | --- | --- | --- | --- | --- |
|  | Mean (ng/mL) | Inter-assay CV (%) | Intra-assay CV (%) | Inter-assay CV (%) | Intra-assay CV (%) |
| Preg | <LLOQ | n/a | n/a | 8.2 | 4.9 |
| Prog | <LLOQ | n/a | n/a | 10.8 | 12.5 |
| DOC | <LLOQ | n/a | n/a | 5.9 | 2.7 |
| B | 2.3 | 11.2 | 7.3 | 12.5 | 3.7 |
| Aldo | <LLOQ | n/a | n/a | 5.6 | 3.7 |
| 17Preg | 4.8 | 16.1 | 16.3 | 14.7 | 8.1 |
| 17OHP | 1.2 | 5.5 | 4.2 | 12.1 | 4.5 |
| S | 0.2 | 8.1 | 6.9 | 5.8 | 3.5 |
| F | 75.2 | 3.5 | 2.4 | 3.7 | 2.5 |
| E | 9.7 | 4.2 | 3.4 | 4.5 | 3.1 |
| DHEA | 2.0 | 6.5 | 6.0 | 6.4 | 6.1 |
| A4 | 0.6 | 15.3 | 8.0 | 9.8 | 9.0 |
| T | 4.0 | 4.7 | 3.5 | 3.2 | 2.3 |
| DHT | 0.4 | 7.5 | 5.4 | 5.4 | 4.1 |
| 5α-dione | <LLOQ | n/a | n/a | 12.8 | 6.1 |
| 3α-adiol | <LLOQ | n/a | n/a | 23.0 | 10.0 |
| An | <LLOQ | n/a | n/a | 4.0 | 2.8 |
| 11KA4 | 0.2 | 16.0 | 16.2 | 8.0 | 6.0 |
| 11KT | 0.1 | 10.1 | 5.9 | 4.7 | 3.0 |
| 11OHA4 | 1.3 | 12.7 | 10.2 | 9.5 | 5.1 |
| 11OHT | 0.1 | 14.9 | 8.2 | 5.3 | 3.2 |
| 5αDHP | <LLOQ | n/a | n/a | 22 | 4.1 |
| alloP | <LLOQ | n/a | n/a | 8.1 | 6.1 |
| 17OHDHP | <LLOQ | n/a | n/a | 17.2 | 7.0 |
| 3α5α17HP | <LLOQ | n/a | n/a | 23.9 | 4.2 |

### 3.4 Serum steroid profiling in healthy volunteers

Following validation, we applied this assay to the measurement of serum samples from 10 female (aged 23-39 years) and 10 male healthy volunteers (aged 28-37 years). The highest concentrations were observed for glucocorticoids (F and E), the adrenal androgen precursor DHEA and the mineralocorticoid precursor B. As expected, men had higher levels of T and DHT than women and the levels of all other adrenal derived steroids were similar in men and women (Figure 3). Concentrations of progesterone and its metabolites 5αDHP and alloP were below the LLOQ in men and in 8 of 10 women. 17OHDHP was not detected in any of the samples (Figure 3). The levels of 17Preg quantified in the samples using our assay were notably higher than expected based on our previous reference ranges and publications by other laboratories. While our validation demonstrates that 17Preg can be measured with acceptable precision (Table 4, 5), we conclude that using the assay presented here 17Preg cannot be quantified accurately in serum.

**Figure 3:**
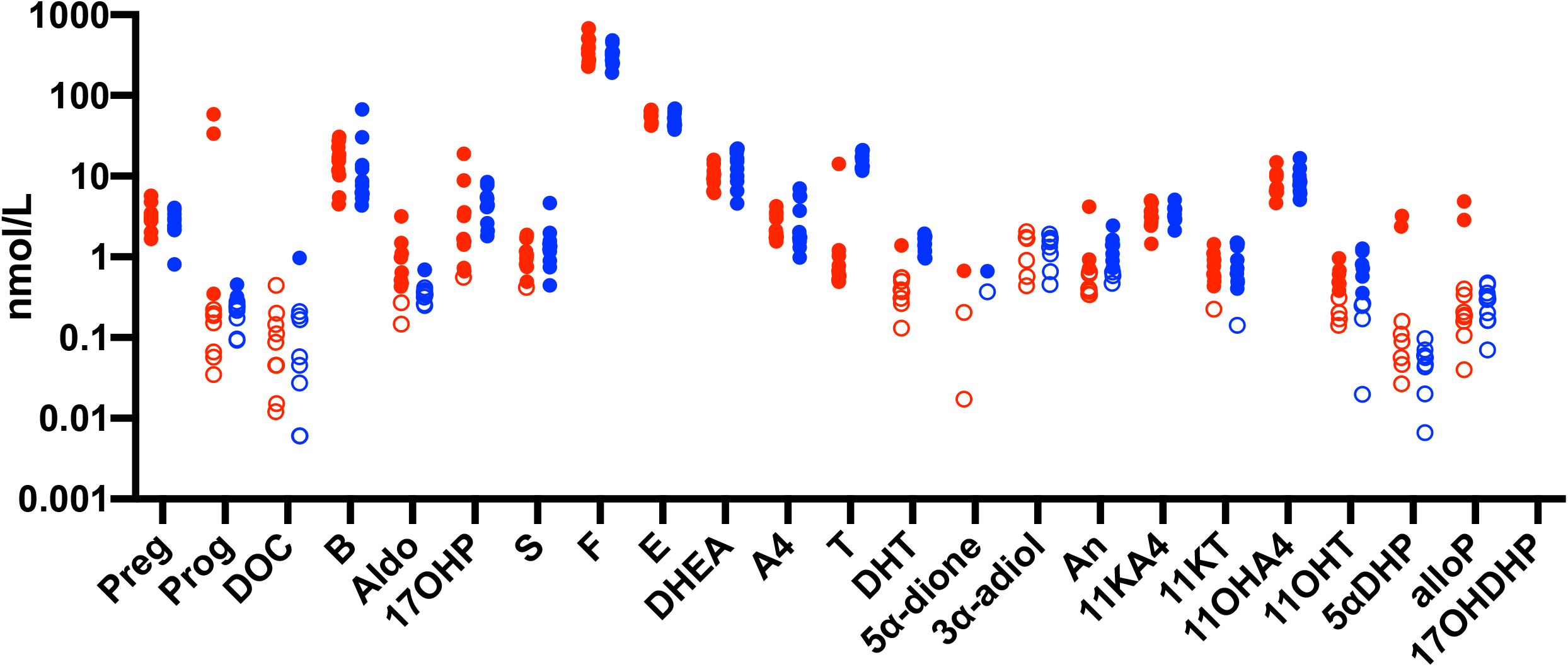
Human Serum Steroid Profiles. Serum steroid profiles in healthy volunteers, female (red circles; n=10, age range 20-40 years) and male (blue circles, n=10, age range 30-40 years). Each symbol represents one participant; values <LLOQ are represented by open circles and undetectable levels were not included in the visualisation. 17OHDHP was undetectable in any of the samples.

## 4. Discussion

Multi-steroid profiling provides a wider insight than quantification of a single steroid marker. A multiplexed approach has extensive utility for the diagnosis, monitoring and the development of a mechanistic understanding of steroid-associated conditions as it allows for the integration of steroid flux across all pathways of steroid biosynthesis and metabolism [2, 25]. This has led several clinical laboratories to increase the number of analytes that are multiplexed in a single assay [14, 26-28]. Here, we describe the development and validation of an uPLC-MS/MS assay for the simultaneous measurement of 25 steroids from the mineralocorticoid, glucocorticoid, and androgen pathways of steroid biosynthesis with a total run-time of 5.5 minutes.

Formic acid is a standard mobile phase additive for the analysis of steroids in positive ionisation mode as 3-keto-Δ4 steroids predominantly form protonated molecular ions [M+H]^+^. However, the sensitivity achieved with formic acid as an additive can still be insufficient to accurately quantify the low levels of certain steroids present in serum. Hence, we tested the effect of post-column infusion of NH_4_F on the signal of all 25 steroids in our assay and established large increases in peak area for all 3-keto-Δ4 analytes in the presence of NH_4_F compared to formic acid, with lower impact observed on the ionisation of A-ring reduced and 3βOH-Δ5 steroids. These findings are consistent with published results on the effect of NH_4_F on the ionisation of selected steroids in a supercritical fluid chromatography set up with ESI in positive mode [19]. 3-keto-Δ4 steroids predominantly form quasi molecular ions [M+H]^+^ during ESI. Parr et al. [19] speculate that the signal enhancement observed for 3-keto-Δ4 is due to the formation of [M+H]^+^ ions being aided in the presence of NH_4_F, as the proton affinity of 3-keto-Δ4 steroids is higher than that of ammonia. Another possible mechanism is that F^-^ ions capture potential Na^+^ contaminations in the mobile phases, hence preventing the formation of [M+Na]^+^ adducts.

While other assays for steroid analysis by ESI in positive mode use NH_4_F as mobile phase additives [18, 29], we chose to supply NH_4_F by post-column infusion to allow for cost- and time efficient incorporation of the assay into lab workflows. As indicated by the system pressure increased in our own preliminary experiments with NH_4_F in the mobile phase, NH_4_F can have detrimental effects on column lifetime [30], which is circumvented by post-column infusion. If running different assays on the same LC-MS/MS system, post-column infusion of additives limits the variability of mobile phase compositions that are used on the system, which could reduce the time needed for equilibration, when switching between assays that require different additives. The use of the in-built fluidics system of the mass spectrometer for the post-column infusion is simple, robust, and fully automated.

The analytical validation of the assay revealed that 23/25 of the analytes can be robustly quantified at typical serum concentrations, while one of the intermediates of the alternative DHT biosynthesis pathway (3α5α17HP) and the precursor steroid 17Preg can only be semi-quantitatively profiled, although the clinical utility of these steroids has yet to be determined. We applied the assay to serum samples from healthy male and female volunteers thereby demonstrating the utility of the assay.

To the best of our knowledge, only two LC-MS/MS assays, which analyse 20 or more steroids in serum or plasma, have been published to date [31, 32]. However, the run-times of these assays (16 minutes [31] and 8 minutes [32]) are significantly longer than that of our assay. Published assays with run-times <6 minutes cover a maximum of 16 analytes [33-36]. Additionally, while other assays focus mainly on adrenal steroids [28, 33, 37-39] or androgen panels [14, 40], our assay comprehensively covers multiple steroid classes (Figure 1). In addition to classic androgens, such as A4 and T, our assay covers the alternative pathway of DHT biosynthesis, which contributes to androgen excess in CAH [41, 42], and the 11-oxygenated androgens, which have only recently been shown to have major relevance for the diagnosis and mechanistic understanding of androgen excess conditions [6, 43-45]. Here we explored the application of this method to interrogate the complex matrix of serum. With minimal adaptations this uPLC-MS/MS method can be applied to other matrices such as bio-fluids (saliva, follicular fluid, micro-dialysis fluid etc.), tissues (adipose, brain, prostate etc.), and *in-vitro* cell/organ culture experiments. These applications would need further validation to determine extraction recovery and matrix effects, but depending on the matrix they could conceivably provide improved accuracy and precision especially at the LLOQ. Our assay has limitations that might be considered to hamper its use in routine clinical laboratories: It uses liquid-liquid extraction, which is inexpensive, but time-consuming and labour-intense, when performed manually, and can be challenging to automate. However, the assay can be adjusted for an extraction technique with higher throughput and better potential for (semi-)automation like supported liquid extraction or C_18_ solid phase extraction, used with satisfying results by other serum multi-steroid assays [14, 28, 39]. Moreover, post-column infusion as a mode of additive delivery is not available on all mass spectrometers, where this is the case external pumps can be purchased to enable post-column infusion. Finally, not all analytes in our assay have their own stable isotope-labelled internal standard, which is usually considered a prerequisite to appropriately control for matrix effects and extraction losses and to allow for accurate and precise quantification [27, 46]. At the time this assay was developed, no internal standards were commercially available for 5α-dione, 11OHT, 5αDHP, 17OHDHP or 3α5α17HP. Therefore, quantification was performed using the internal standard eluting closest to the analyte, our validation data proves this approach to be sufficient. Use of customised deuterium- or ^13^C-labelled internal standards is expensive but could improve accuracy and imprecision of these analytes.

## 7. Conclusions

uPLC-MS/MS with post-column infusion of ammonium fluoride enables the high throughput, sensitive profiling of 25 steroids. Use of NH_4_F significantly increased sensitivity for most steroids. The method was analytically validated and applied to human serum.

## Data Availability

All data produced in the present study are available upon reasonable request to the authors

## Acknowledgments

The authors thank the volunteers for the donation of blood samples.

## Research funding

Wellcome Trust (Investigator Award WT209492/Z/17/Z, to W.A.) Academy of Medical Sciences UK (Newton Advanced Fellowship NAF004\1002, to K.H.S).

