## Supplemental tables and figures for "Multi-steroid profiling by uPLC-MS/MS with post-column infusion of ammonium fluoride"

**Supplemental Table 1: Suppliers of standards and stable isotope labelled internal standards.**

| **Steroid abbreviation** | **Steroid trivial name** | **Supplier of standard and catalogue #** | **Internal standard used for quantification** | **Supplier of internal standard**  **and catalogue #** |
| --- | --- | --- | --- | --- |
| Preg | Pregnenolone | Sigma-Aldrich  P9129 | Preg-d2-^13^C3 | Sigma-Aldrich  P109 |
| Prog | Progesterone | Sigma-Aldrich  P8783 | Prog-d9 | Cambridge Isotope Labs  DLM-7953 |
| DOC | 11-deoxycorticosterone | Sigma-Aldrich  D6875 | DOC-d8 | Steraloids  Q3460-020 |
| B | Corticosterone | Sigma-Aldrich  C2505 | B-d8 | Cambridge Isotope Labs  DLM-7347 |
| Aldo | Aldosterone | Steraloids  Q2000-000 | Aldo-d7 | IsoSciences  55093 |
| 17Preg | 17α-hydroxypregnenolone | Sigma-Aldrich  H5002 | 17OHPreg-d3 | CDN Toronto Research Chemicals  D5329 |
| 17OHP | 17α-hydroxyprogesterone | Sigma-Aldrich  H5752 | 17OHP-d8 | Cambridge Isotope Labs  DLM-6598 |
| S | 11-deoxycortisol | Sigma-Aldrich  R0500 | S-d2 | Cambridge Isotope Labs  DLM-7209 |
| F | Cortisol | Sigma-Aldrich  H4001 | F-d4 | Cambridge Isotope Labs  DLM- 2218 |
| E | Cortisone | Sigma-Aldrich  C2755 | E-d7 | Sigma Aldrich 705586 |
| DHEA | Dehydroepiandrosterone | Sigma-Aldrich  D4000 | DHEA-d6 | Sigma-Aldrich  709549 |
| A4 | Androstenedione | Sigma-Aldrich  46033 | A4-d7 | CDN Toronto Research Chemicals D5305 |
| T | Testosterone | Sigma-Aldrich  T6147 | Test-d3 | Sigma-Aldrich  T2655 |
| DHT | 5α-dihydrotestosterone | Sigma-Aldrich  A8380 | DHT-d3 | Cerilliant  D077 |
| 5α-dione | 5α-androstanedione | Steraloids  A1630 | DHT-d3 | Cerilliant  D077 |
| 3α-adiol | 5α-androstanediol | Sigma-Aldrich  A7755 | 3α-adiol-d3 | CDN Toronto Research Chemicals  A637527 |
| An | 5α-androsterone | Steraloids  A2420 | An-d4 | IsoSciences  14235 |
| 11KA4 | 11-ketoandrostenedione | Sigma-Aldrich  284998 | 11KA4-d10 | Toronto Research Chemicals A305604 |
| 11KT | 11-ketotestosterone | Steraloids  A6720 | 11KT-d3 | IsoSciences  16144 |
| 11OHA4 | 11β-hydroxyandrostenedione | Sigma-Aldrich  A3009 | 11OHA4-d7 | Cambridge Isotope Labs  DLM-9697 |
| 11OHT | 11β-hydroxytestosterone | Steraloids  A5760 | 11OHA4-d7 | Cambridge Isotope Labs  DLM-9697 |
| 5αDHP | 5α-dihydroprogesterone | Sigma-Aldrich  P7754 | 5αDHP-d4 | Steraloids  P3830 |
| alloP | Allopregnanolone | Steraloids  P3800 | alloP-d4 | Steraloids  P3830 |
| 17OHDHP | 17α-hydroxy-5α-dihydroprogesterone | Steraloids  3700 | DHT-d3 | Cerilliant  D077 |
| 3α5α17HP | 17α-hydroxyallopregnanolone | Steraloids  P2460 | An-d4 | IsoSciences  14235 |

**Supplemental Table 2: Concentrations of the calibrators and quality controls (QC) for each analyte in ng/mL and approximate concentration in nmol/L calculated for a molecular weight of 300.**

| **Name** | **Concentration**  **(ng/mL)** | **Approximate concentration**  **(nmol/L)** |
| --- | --- | --- |
| C0 | 0 | 0 |
| C1 | 0.05 | 0.2 |
| C2 | 0.1 | 0.3 |
| C3 | 0.5 | 1.6 |
| C4 | 1 | 3.3 |
| C5 | 2.5 | 8.3 |
| C6 | 5 | 16.6 |
| C7 | 10 | 33.3 |
| C8 | 25 | 83.3 |
| C9 | 50 | 166.6 |
| C10* | 100 | 333.3 |
| C11*^+^ | 250 | 833.3 |
| QC 1 | 0.3 | 1 |
| QC 2 | 1 | 3.3 |
| QC 3 | 3 | 10 |
| QC 4 | 30 | 100 |

* excluded for A4 and T; ^+^ excluded for Prog, DOC, B, Aldo, 17Preg, 17OHP, S, F, E, DHEA, A4, T, DHT, 5α-dione, An, 11KA4, 11KT, 11OHA4, 11OHT, 5αDHP to avoid saturation of the mass spectrometer

**Supplemental Figures**

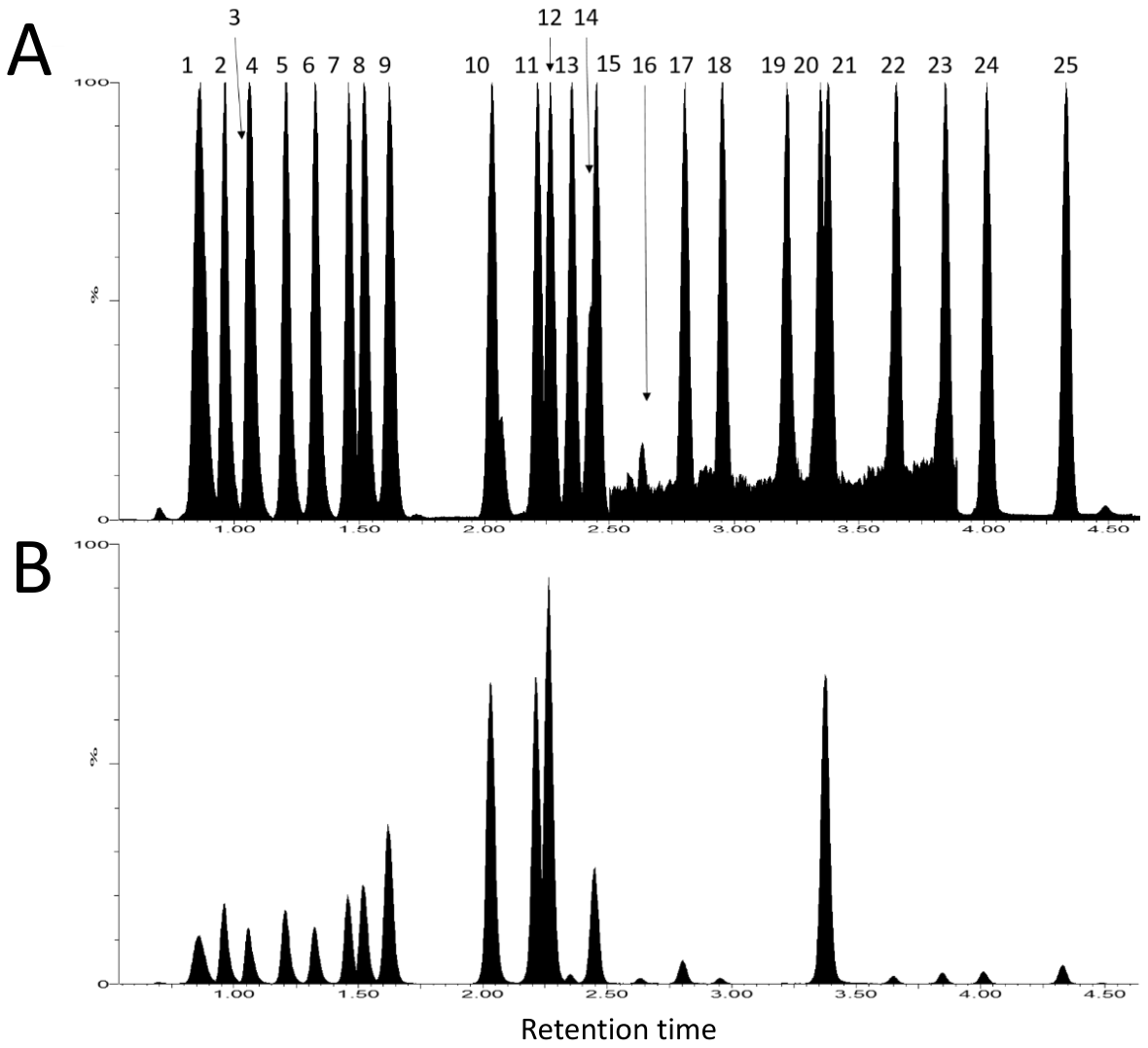

**Supplemental Figure 1: Chromatogram of a sample with all target analytes (A) and internal standards (B).** 1: Aldo, 2: E, 3: F, 4: 11KA4, 5: 11KT, 6: 11OHA4, 7: 11OHT, 8: B, 9: S, 10: A4, 11: DOC, 12: T, 13: DHEA, 14: 17OHP, 15: 17Preg, 16: 5α-dione, 17: DHT, 18: 17OHDHP, 19: 3α-adiol, 20: Prog, 21: An, 22: Preg, 23: 3α5α17HP, 24: 5αDHP, 25: alloP

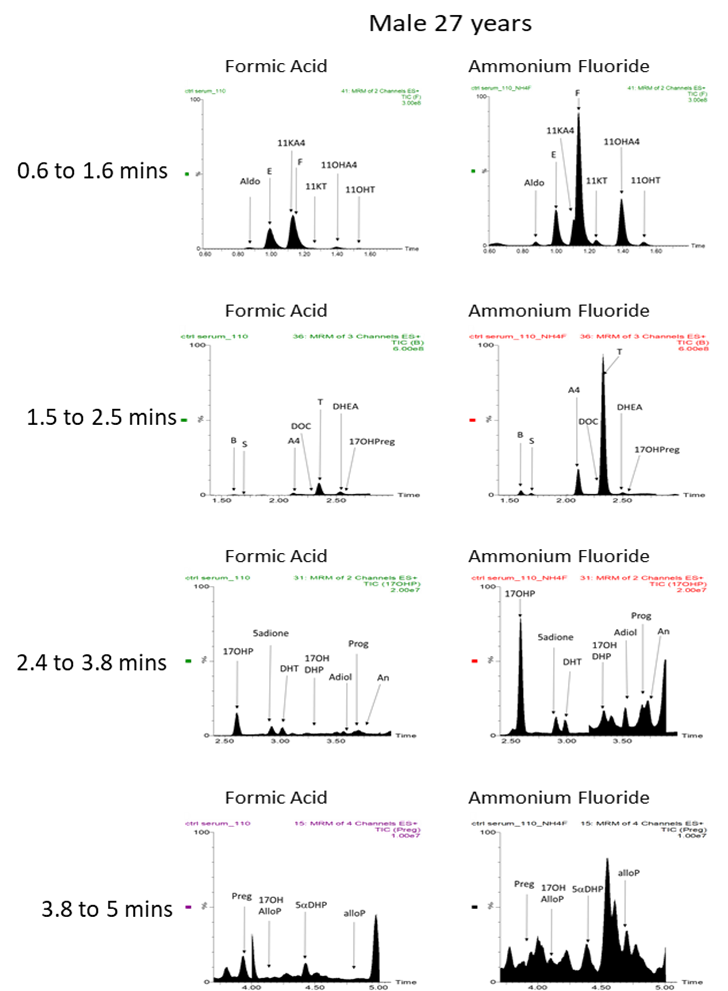

**Supplemental Figure 2: Representative chromatograms for 25 steroids in serum from a male in his late 20’s using formic acid as mobile phase additive (0.1% (v/v) in mobile phase A (water) and B (methanol); left panels) or post-column infusion of NH_4_F (6 mmol/L, 5 μL/min) with 0.1% (v/v) formic acid in mobile phase A (water; right panels).** Traces show the TIC for the respective analyte.
